# Personalising Alzheimer’s Disease progression using brain atrophy markers

**DOI:** 10.1101/2023.06.15.23291418

**Authors:** Serena Verdi, Saige Rutherford, Charlotte Fraza, Duygu Tosun, Andre Altmann, Lars Lau Raket, Jonathan M. Schott, Andre F. Marquand, James H. Cole for the Alzheimer’s Disease Neuroimaging Initiative

## Abstract

INTRODUCTION: Neuroanatomical normative modelling can capture individual variability in Alzheimer’s Disease (AD). We used neuroanatomical normative modelling to track individuals’ disease progression in people with mild cognitive impairment (MCI) and patients with AD.

METHODS: Cortical thickness and subcortical volume neuroanatomical normative models were generated using healthy controls (n∼58k). These models were used to calculate regional Z-scores in 4361 T1-weighted MRI time-series scans. Regions with Z-scores <-1.96 were classified as outliers and mapped on the brain, and also summarised by total outlier count (tOC).

RESULTS: Rate of change in tOC increased in AD and in people with MCI who converted to AD and correlated with multiple non-imaging markers. Moreover, a higher annual rate of change in tOC increased the risk of MCI progression to AD. Brain Z-score maps showed that the hippocampus had the highest rate of atrophy change.

CONCLUSIONS: Individual-level atrophy rates can be tracked by using regional outlier maps and tOC.

## 1 Background

Alzheimer’s Disease (AD) interacts with an individual’s distinct genetic and environmental influences, leading to a unique pattern of brain atrophy that changes dynamically as the disease progresses.^1–4^ This heterogeneity impacts individual differences in the time of symptom onset and the severity and progression of symptoms.^3^ While well-known clinically, this heterogeneity is often overlooked in research studies and clinical trials, creating challenges in the clinic (e.g., when planning treatment and care) and complicating clinical trial recruitment.^5–10^ Therefore, there is a need to measure disease heterogeneity at the individual level.^11^

Neuroimaging provides insights into brain structure in vivo, rendering it an ideal tool for studying AD, yet most studies focus on group-average or subtype effects, overlooking the individual variability between patients.^12, 13^ To overcome this reliance on group averages, neuroanatomical normative modelling is an emerging technique which captures individual-level variability in the brain. ^14, 15^ Based on the well-established normative modelling concept, as per height and weight growth charts for children ^16^, the neuroanatomical version builds separate normative models per brain region, based on a large independent reference dataset. A new individual can then be compared to these normative models to ascertain whether their brain volume or cortical thickness is lesser or greater than would be expected for someone of their age and sex. This deviation from normality can be quantified using Z-scores, of which brain-wide maps of Z-scores can be generated, providing a unique fingerprint of brain health relative to the norm for individual patients.^3^ Therefore, a great advantage of neuroanatomical normative modelling is that it can detect if a brain has been adversely affected by Alzheimer’s in multiple regions. Moreover, as neuroanatomical normative models compare single patient scans to an independent dataset, the patient can be considered individually, providing a natural fit to personalised healthcare and precision medicine.

Recent applications of neuroanatomical normative modelling have generated personalised maps of individual patients’ atrophy patterns. Using neuroimaging data from a large AD research cohort (Alzheimer’s Disease Neuroimaging Initiative (ADNI)) we have previously shown that cortical thinning patterns vary from patient to patient.^17^ Here, individualised brain-wide Z-score maps revealed heterogenous atrophy patterns in the AD group, additionally cortical thinning heterogeneity in people with MCI was predictive of conversion to AD. Furthermore, relationships between cortical thickness heterogeneity and cognitive function, amyloid-beta, phospho-tau, and ApoE genotype were observed. Similarly, atrophy patterns were shown to be related to disease severity, presenting phenotypes and comorbidities in amyloid-positive AD patients in a study using real-world memory clinic data.^18^ These studies corroborate the heterogeneity of brain atrophy in AD and provide evidence that neuroanatomical normative modelling can be used to explore anatomical-clinical correlations at an individual level. However, the insights from these studies were based only on cross-sectional neuroimaging data. To fully understand the potential of neuroimaging biomarkers for clinical use and to comprehend causal mechanisms, longitudinal data is necessary. This will aid in understanding how patterns of atrophy change over time when monitoring disease progression at an individual level.

In this work we applied the neuroanatomical normative modelling technique to quantify regional changes in neuroanatomical variation across the AD disease course. We used an emerging neuroanatomical normative modelling technique, which can map brain ageing at high spatial precision. Moreover, this model can be optimised with the inclusion of controls scanned at the same site of the clinical cohort (adaptive learning).^14^ We assessed whether markers of regional brain atrophy variation could track disease progression in people with MCI and patients with AD. We also explored how the rate of change in atrophy related to other common imaging and non-imaging AD markers.

## Methods

### 2.1 Participants and patient dataset

Participants were derived from two datasets: (1) a reference dataset comprised of healthy people across the human lifespan, and (2) a clinical target dataset which included people with AD or MCI in addition to age-matched cognitively normal controls. The reference dataset was made by combining data on healthy people from multiple publicly available sources ^19^, including Open Access Series of Imaging Studies (OASIS), Adolescent Brain Cognitive Development (ABCD) study and UK Biobank (UKB), totalling 58,836 from 82 sites. Data collection, data processing and participant demographics of the reference dataset were described previously^14^. The clinical data used in the preparation of this article was obtained from the ADNI database; http://adni.loni.usc.edu. Inclusion criteria were the availability of T1-weighted MRI scans (acquired using 1.5T and 3T MRI scanners), which had at least 12 weeks apart between scanning visits. Here data had a maximum range of 9.5 years (115 months). Furthermore, AD participants had to meet the National Institute of Neurological and Communicative Disorders and Stroke–AD and Related Disorders Association criteria for probable AD. MCI participants reported a subjective memory concern either autonomously or via an informant or clinician, participants had no significant levels of impairment in other cognitive domains. Further to this criterion, we labelled participants with MCI as either those that continued to exhibit MCI as ‘MCI stable’, or conversely as those who progressed to being diagnosed with AD in the study period (115 months) as ‘MCI progressive’.

#### 2.1.1 Standard Protocol Approvals, Registrations, and Patient Consents

Written informed consent was obtained from all participants before experimental procedures were performed. Approval was received by an ethical standards committee for ADNI study data use.

### 2.2 MRI acquisition

For the clinical dataset, T1-weighted images were acquired at multiple sites using 1.5T or 3T MRI scanners. Detailed MRI protocols for T1-weighted sequences are available online (http://adni.loni.usc.edu/methods/documents/mri-protocols/). The quality of raw scans was evaluated at Mayo Clinic for technical problems and significant motion artefacts and clinical abnormalities. ^20^

### 2.3 Estimation of cortical thickness and subcortical volumes

T1-weighted scans from both the reference and the clinical dataset underwent processing using automated FreeSurfer to extract the cortical thickness of 148 cortical regions and grey matter tissue volume of 20 subcortical volumes from the Destrieux parcellations.^21, 22^ For both datasets a mix of version 5 and 6 of FreeSurfer was used. Quality control of FreeSurfer processing for the reference dataset relied on both manual and automated filtering, as described previously.^14^ For the clinical dataset, quality control was based on a visual review of each cortical region performed at UCSF (https://adni.bitbucket.io/reference/docs/UCSFFSX51/UCSF%20FreeSurfer%20Methods%20and%20QC_OFFICIAL.pdf).

### 2.4 Neuroanatomical Normative modelling

A non-Gaussian Bayesian Regression model was implemented, which accounts for the non-Gaussian nature of MRI data and adjusts for unwanted noise from scanning acquisition across multiple sites.^23^ This model was trained on multi-site data to generate normative models per region using the covariates age and sex and site. Here, by training on 58,836 scans from datasets across 82 sites, the model produces a stable distribution of estimates across the entire lifespan – details of this process have been previously outlined.^14, 23^ Next, these estimates were conditioned to our specific context, using an adapted transfer learning approach.^19^ The parameters of the reference normative model were recalibrated to the longitudinal ADNI dataset using 70% of cognitively normal healthy controls per ADNI site, where 70% was used to give stable estimates of the transferred model parameters, given that many of the scan sites in ADNI have small sample sizes. The remaining 30% of healthy controls, plus people with MCI and patients with AD were used for subsequent analysis, with Z-scores generated per region for each scan. The modelling steps and processed reference dataset are openly available: https://github.com/predictive-clinical-neuroscience/braincharts.

#### 2.4.1 Individualised brain markers

Outliers in terms of low cortical thickness were identified for each region, defined as Z < - 1.96 (corresponding to the bottom 2.5% of the normative distribution of cortical thickness). The number of outliers were summed across 168 regions for each participant, to give a total outlier count (tOC) across regions. Brain surface mapping was conducted using the Destrieux (148 cortical regions) and aseg (19 subcortical regions) atlas via the R package *ggseg.* All statistical analyses were implemented in R version 3.6.2.

### 2.5 Disease course analysis

#### 2.5.1 Yearly timepoint assessment

For these analysis, longitudinal data were subset into three time-points representing a cross-sectional measure of baseline, 12-month and 24-month of study visits. Here study visits between 36 – 115 months was excluded because of a low number of study participants at each time point due to study attrition. Brain outlier maps for each diagnostic group were mapped across the three time points. This enabled visualisation of the extent to which patterns of outlier regions overlap or are distinct across three years.

#### 2.5.2 Rate of change in tOC and regional Z-scores

To assess longitudinal atrophy, we took two related approaches. First, we calculated the rate of change in tOC as the difference in baseline and final tOC (up to 115 months). A linear mixed-effect model was used to test for differences in the rate of change in tOC between diagnostic groups, whilst adjusting for age, sex and predicted Alzheimer’s disease progression (AC score, which represents predicted years since amyloid PET positivity).^24, 25^ Second, we calculated the rate of change in Z-score per region. Here, we calculated the difference between the baseline and final Z-score, then we defined a new ‘normative model’ based on the distribution of rates of change in scans of cognitively unimpaired controls (n = 610). We then classified individuals as ‘rate of Z-score change outliers’, if their rate of change was more than two standard deviations away from the mean in the controls (which was Z = -0.0009). Then, the neuroanatomical patterns of the ‘rate of Z-score change outliers’ were mapped onto brain surfaces for visualisation purposes. As a final step, to provide more detail, we focused on the region of the highest rate of change (in this case, the left hippocampus), and compared patients with AD who were ‘rate of Z-score change outliers’ with those that were not, based on their total Mini-Mental State Examination (MMSE) score and CSF Aβ 1-42 and p-tau181 values.

#### 2.5.3 Growth models

Multilevel growth models were used to predict the trajectory of tOC over time using the lme4 package in R. Here, a linear mixed effects model was generated, with a random effect of participant (including slope and intercept), and a fixed-effects interaction between time and group, while covarying for age and sex.

#### 2.5.4 MCI to AD conversion analysis

Follow-up diagnosis status data, up to three years from the baseline scan, were obtained from 647 people with MCI. We ran a survival analysis using Cox proportional hazards regression to assess whether tOC related to the risk of converting from MCI to AD in 3 years (MCI progressive n=75). This included assessing 1-year change in tOC (i.e., the difference in tOC between baseline and month 12) as a predictor variable to explore how the change in tOC is associated with disease progression. We also compared the 1-year change in tOC between MCI stable and MCI progressive groups.

### 2.6 Relationship to other disease markers

#### 2.6.1 Alzheimer’s staging score

An Alzheimer’s disease staging score based, representing predicted years since amyloid PET positivity, was calculated based on latent-time disease-progression modelling of longitudinal amyloid PET SUV_R_ and clinical scores (CDR-SB, ADAS-cog, MMSE).^24, 26^ This disease staging score (AC score) has previously been shown to better stage patients along the Alzheimer’s continuum than conventionally used staging measures (e.g., early or late MCI) ^24^ – this continuous staged measure was used as a covariate to adjust for disease stage when exploring how tOC relates to cognitive, amyloid and tau AD disease markers.

#### 2.6.2 Cognitive markers

Linear regression adjusting for age, sex and years of education examined the relationship between tOC rate of change and cognitive composite scores (memory using ADNI MEM or executive function using ADNI EF).^34^ We then assessed the interaction between the diagnostic group and cognitive composite score. Total MMSE scores were only used in disease conversion and regional rate of change comparisons. The collection date of the cognitive variables were matched (within a 12-week range) to the date of TI-MRI acquisition.

#### 2.6.3 Amyloid and Tau markers

Florbetapir and flortaucipir were used to index cerebral amyloid and tau deposition. For tau, a summary SUV_R_ was generated by taking the mean across all brain regions and dividing by inferior cerebellar GM (reference region) as detailed previously (https://adni.bitbucket.io/reference/docs/UCBERKELEYAV1451/UCBERKELEY_AV1451_Methods_Aug2018.pdf). This summary SUV_R_ which was matched to 133 longitudinal T1-weighted MRI scans (total of 107 participants). For amyloid, summary SUV_R_ was based on the whole cerebellum reference region for 667 cases with matching T1-weighted MRI scans (total of 489 participants). Details of the PET processing methods are provided by Landau et.al, UC Berkeley, and can be found on the ADNI database; http://adni.loni.usc.edu.27 Both measures were matched from the PET scanning date to the MRI T1-weighted date scan date in a 12-week range. CSF amyloid-beta (Aβ 1-42) and phospho-tau 181 were also employed as amyloid and tau markers in this study, which were matched (within a 12-week range) to the date of TI-MRI acquisition. CSF amyloid had a total of 858 cases matched (n=294), and CSF phospho-tau 181 had a total of 856 cases (n= 293). A linear regression adjusting for age and sex examined the relationship between the rate of change in tOC and amyloid and tau markers. We assessed the interaction between the diagnostic group and amyloid and tau markers in a subsequent regression.

#### 2.6.4 Genetic markers

ApoE ε4 status was determined by either being ApoE ε4 homozygous, ApoE ε4 heterozygous or ApoE ε4 non-carrier. Group differences in the rate of change in tOC were assessed as a linear regression, adjusting for age and sex. A Polygenic Risk Score (PRS) was previously generated for all ADNI participants with genome-wide data by Altmann and colleagues.^28^ Effects for APOE-ε2 and APOE-ε4 were manually added using effect sizes. A p-value inclusion cut-off was used which included SNPs passing genome-wide suggestive significance (*P* = 1.0×10^-05^). The relationship between PRS and rate of change in tOC was examined using linear regression, adjusting for age, sex and ApoE ε4 status. We assessed the interaction between the diagnostic group and PRS in a subsequent regression.

## 3 Results

### 3.1 Participants

A total of 1,492 participants and 4,361 scans from across 62 ADNI sites were included (**Table 1**). Here 70% of controls were removed from the clinical dataset and were used as a calibration dataset to adapt the normative model to the new sites. These were randomly selected and stratified across sites and gender to make sure all sites and genders are present in the adaptation set. The final clinical dataset amounted to a total of 1181 participants with a total of 3362 scans.

**Table 1.** Demographics of the clinical sample (ADNI), Key: * adjusting for age, sex and ApoE ε4, ** *X*^2^ of ApoE ε4 status. ^†^ Disease stage score represents predicted years since amyloid PET positivity

|  | Controls | MCI | AD | Total | Statistical differences |
| --- | --- | --- | --- | --- | --- |
| **n** | 245 | 647 | 245 | 1181 | - |
| **total of scans** | 610 | 2095 | 595 | 3362 | - |
| **Median (IQR) of scans per participant** | 3(2) | 4(3) | 3(2) | 4(3) | - |
| **Follow-up time in time series data (months) mean ± sd** | 37.4 ± 25.8 | 30.6 ± 24.8 | 13.9 ± 12.01 | 29.3 ± 24.5 | $F(2, 603)= 34.4$,<br>$P=9.6 \times 10^{-15}$ |
| **Sex (M:F)** | 110:159 | 289:236 | 106:100 | 505:495 | $X^2= 14.32$,<br>$P=7.6 \times 10^{-04}$ |
| **Baseline age mean ± sd & range** | 74.8 ± 6.4<br>(57-93) | 73.1±7.7<br>(55-92) | 74.4 ± 8.1<br>(55-91) | 73.8 ± 7.5<br>(55-93) | $F(2, 997)=4.78$,<br>$P=0.008$ |
| **Baseline AC score (years)<sup>†</sup> mean ± sd & range** | -4.55 ± 5.8<br>(-15.3 – 9.9) | 7.23 ± 3.8<br>(-3.5 – 15.8) | 13.3 ± 2.1<br>(2.5-16.5) | 4.37 ± 8.52<br>(-15.3 – 16.5) | $F(2, 607)=868.4$,<br>$P=2.2 \times 10^{-16}$ |
| **MMSE score mean ± sd total & range** | 28.95 ± 1.31<br>(22-30) | 27.78 ± 2.03<br>(19-30) | 22.05 ± 3.77<br>(5-30) | 26.89 ± 3.47<br>(5-30) | $F(2, 845)=486.8$,<br>$P=2.2 \times 10^{-16}$ |
| **Baseline ADNI memory mean ± sd total & range** | 1.23 ± 0.62<br>(-0.3- 3.3) | 0.39 ± 0.7<br>(-1.6-2.4) | - 0.9 ± 06<br>(-2.87- 0.39) | 0.35 ± 1.01<br>(-2.8-3.3) | $F(2, 891)=604.8$,<br>$P=2.2 \times 10^{-16}$ |
| **Baseline ADNI executive function mean ± sd total & range** | 1.06 ± 0.81<br>(-0.9-2.9) | 0.36 ± 0.87<br>(-1.9-2.9) | -0.99 ± 1.06<br>(-3.0-2.6) | 0.28 ± 1.14<br>(-3.0-2.9) | $F(2, 886)=279.6$,<br>$P=2.2 \times 10^{-16}$ |
| **Baseline CSF Aβ 1-42 (pg/mL) mean ± sd total & range** | 335.46 ± 386.9<br>(90.7 - 2099) | 337.89 ± 440<br>(82.5- 2501) | 200 ± 215.6<br>(80.4- 1286) | 300.83 ± 385.52<br>(80.4- 2501) | $F(2, 291)=3.69$,<br>$P=0.03$ |
| **Baseline CSF p-tau (pg/mL) mean ± sd total & range** | 33.07 ± 18.5<br>(8.8-98.5) | 38.19 ± 23.0<br>(8.53- 121) | 58.02 ± 34.3<br>(12.8- 188) | 42.39 ± 27.38<br>(8.53-188) | $F(2, 290)=20.1$,<br>$P=6.6 \times 10^{-09}$ |
| **Baseline amyloid PET SUVR mean ± sd total & range** | 1.12 ± 0.23<br>(0.85-2.69) | 1.19 ± 0.22<br>(0.18- 0.84) | 1.44 ± 0.19<br>(0.93- 1.82) | 1.21 ± 0.24<br>(0.18-2.26) | $F(2, 486)=61.0$,<br>$P=2.2 \times 10^{-16}$ |
| **Baseline tau PET SUVR mean ± sd total & range** | 1.50 ± 0.18<br>(1.15-2.04) | 1.56 ± 0.23<br>(1.21- 2.41) | 1.84 ± 0.42<br>(1.48- 2.75) | 1.56 ± 0.25<br>(1.15-2.75) | $F(2, 104)=10.6$,<br>$P=6.2 \times 10^{-05}$ |
| **ApoE ε4 non-carrier (proportion in group sample)** | 69.0% | 57.1% | 31.2% | 55.3% | $**X^2= 90.41$,<br>$P=2.2 \times 10^{-16}$ |

### 3.2 The change in neuroanatomical outliers between baseline and 24 months

The proportion of cortical thickness and subcortical volume outliers defined in each group differed in regional patterns between Alzheimer’s, MCI and control groups, across 24 months (**Fig. 1**). Here the proportion of outliers in each diagnostic group was mapped cross-sectionally at yearly time points over 24 months. In patients with AD, we found that the proportion of outliers differed with each yearly time point. Here, the region with the highest proportion of outliers is consistently the left hippocampus, with 47% at baseline, 60% at 12 months and 72% at 24 months. Yet, the total number of cortical and subcortical outliers fluctuated with no particular trend; at baseline there were 134 cortical regions and 12 subcortical regions with outliers, to 128 and 9 at 12 months and 131 and 11 at 24 months respectively.

**Fig 1.**
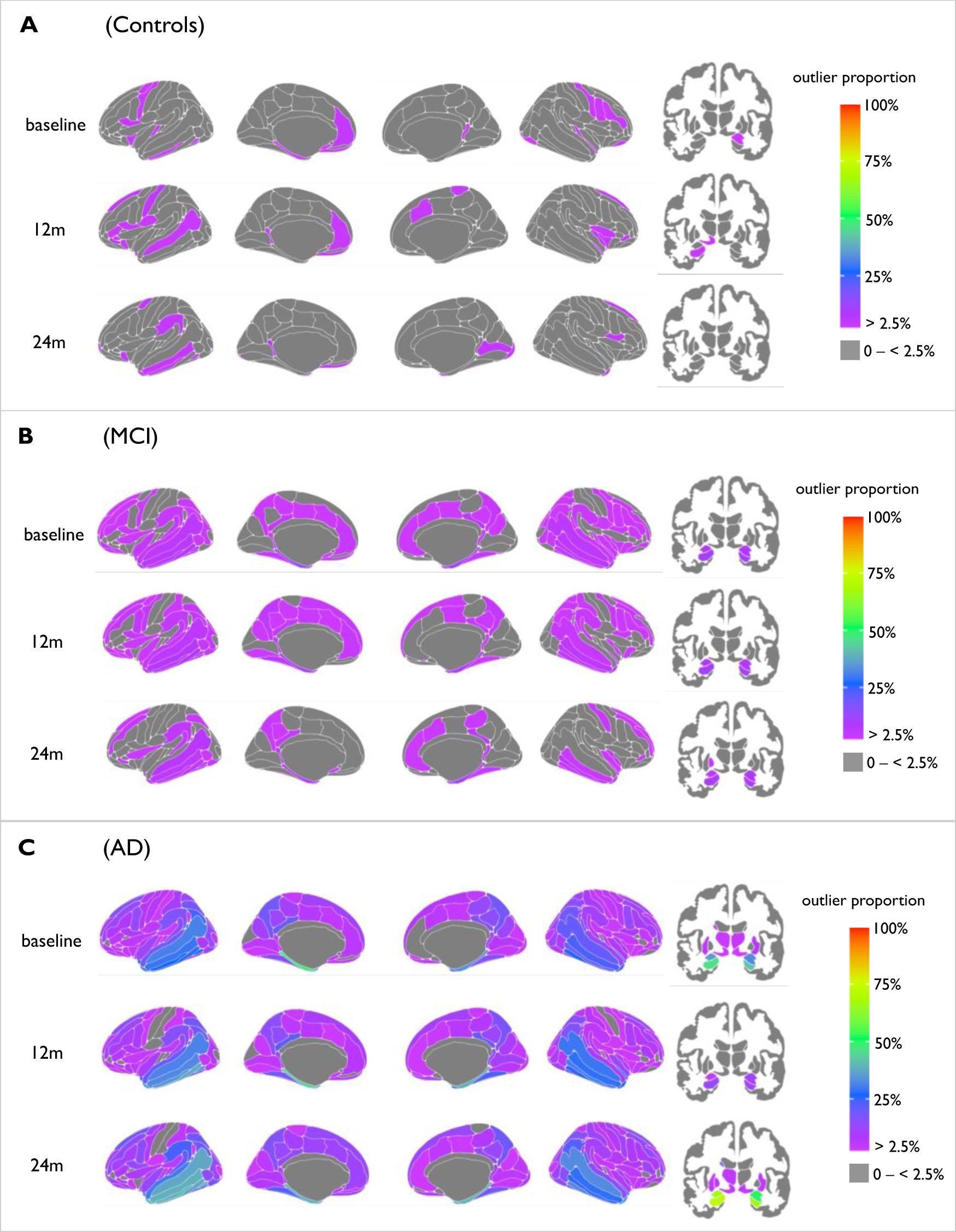
Mapped are the percentage of outliers present in each diagnostic group at 1^st^ – 3^rd^ year of visits, for both cortical (left) and subcortical (right) areas. The colour bar reflects outlier proportion from 2.5% to 100% (thresholding of Z-scores). Zero percent (grey) represents that no participants have outliers in those respective regions.

### 3.3 Higher rate of atrophy in brain regions in patients with AD

In subcortical areas the highest proportion of rate of change was in the left hippocampus which was 52%, therefore 48% of AD patients did not follow this trend of increased atrophy here (**Fig. 2**). When comparing patients with AD that did or did not have heightened atrophy rates there was no difference in total MMSE score, AC score, p-tau [P>0.05]. There was a difference in CSF Aβ 1-42 [*β* = 80.37, *P* = 0.0003], here patients with AD that had heightened atrophy rate in the left hippocampus had higher CSF Aβ 1-42 levels, as (mean = 209.98, sd = 224.87) compared to those did not (mean = 129.61, sd = 30.02).

**Fig 2.**
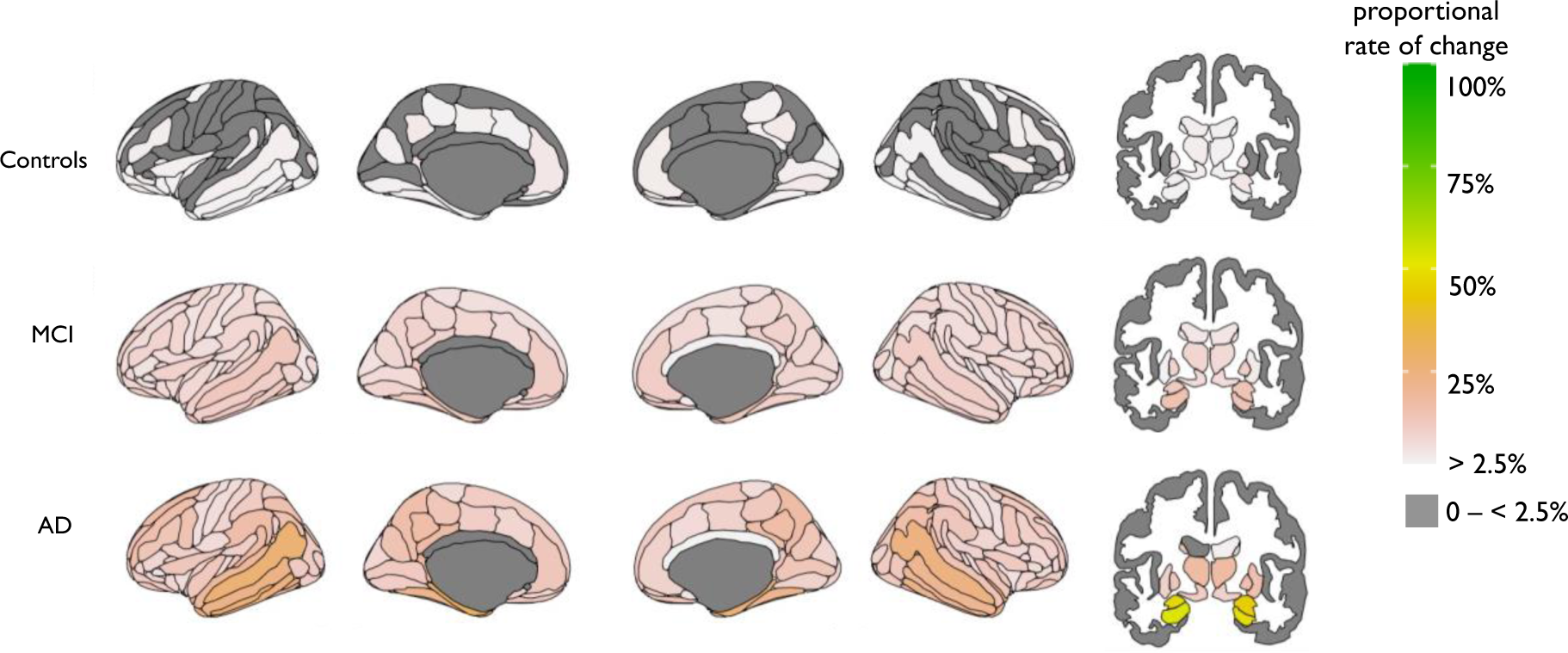
Mapped is the proportion of participants with a high rate of atrophy in cortical (left) and subcortical (right) areas.

### 3.4 tOC increases with time in AD and in people with MCI who convert to AD

Growth models showed an increase in tOC over time was observed in the AD group (β = 0.38, P = 6.8×10^-12^) and in the MCI group (β = 0.01, P = 0.002) but not in controls (β = - 0.001, P = 0.79) (**Fig 3, panels A;B**). Furthermore, linear regression revealed that there was a significant increase in the rate of tOC change over time that differed between groups overall when adjusting for baseline AC score, baseline age and sex [F(5, 382) = 7.07, P = 2.4×10^-06^]. Pairwise comparisons (Tukey post-hoc) were significant [P<=0.001] in AD versus controls and AD versus MCI, however, this was not significant in controls versus MCI [P= 0.93], with the rate of change in tOC being highest in the AD group (mean increase = 5.43, SD=14.19), intermediate in the MCI group (mean=0.31, SD= 5.54) and lowest in controls (mean = 0.06, SD= 1.88) (**Fig 3, panel C**). Further to these diagnostic groups, the severity of dementia reflected by the AC score was significantly associated with the rate of change in tOC across the whole sample, when adjusting for age and sex (β = 0.22, P = 3.7×10^-05^).

**Fig. 3.**
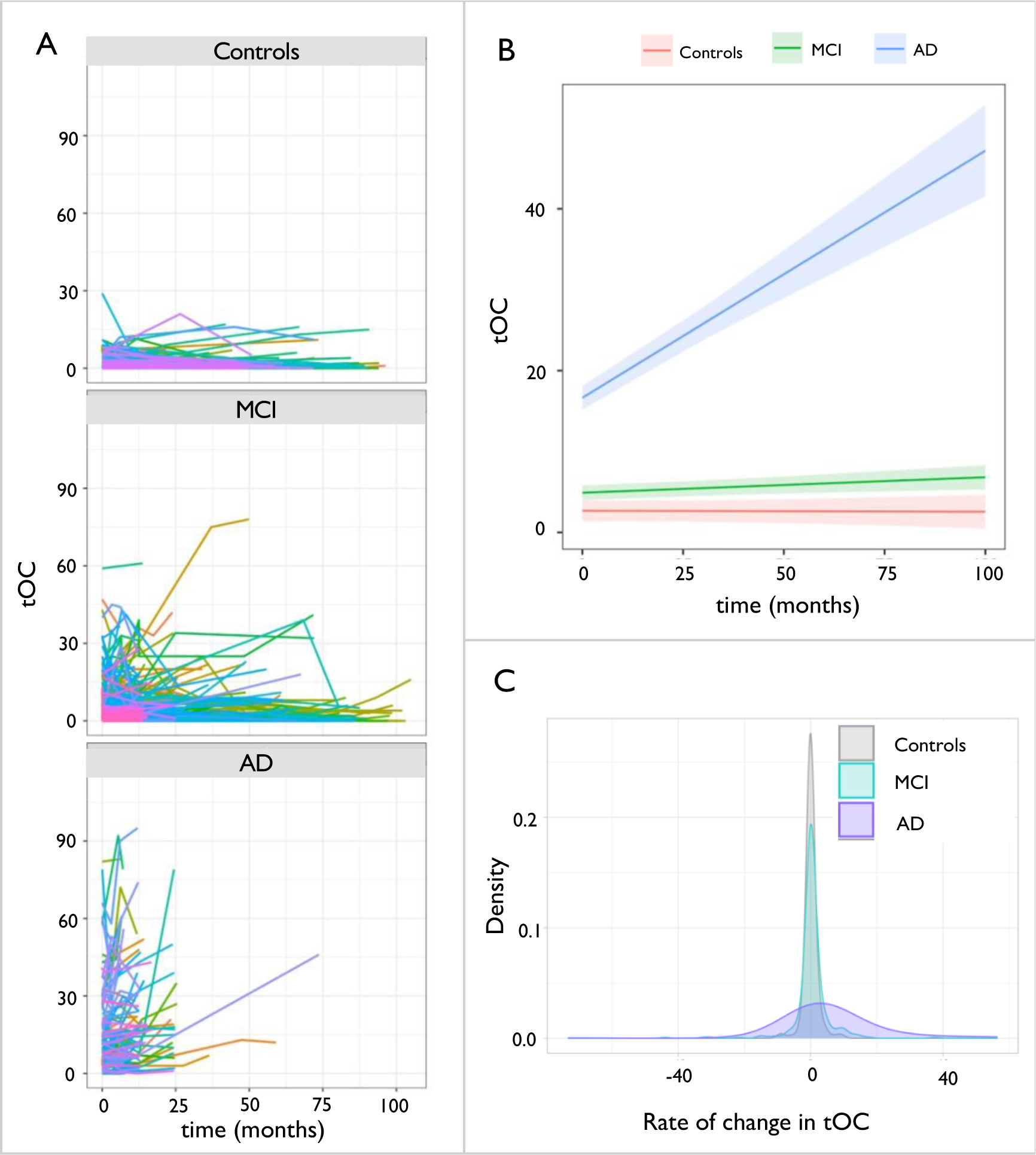
Change in total outlier count according to diagnostic group. **(a)** Spaghetti plot of tOC according to diagnostic group. Each line represents an individual participants trajectory of tOC scores over the scanning period **(b)** Linear growth model for each diagnostic group **(c)** Density plot of the rate of change in tOC in diagnostic groups. Across the whole sample the rate of change ranged from -73.18 to 56.29, mean = 1.16, SD= 7.6.

Growth models showed that an increase in tOC over time was observed in both the MCI progressive group (β = 0.10, P = 4.8×10^-05^) and the MCI stable group to a lesser extent (β = 0.08, P = 0.002) (**Fig 4, panels A;B**). Furthermore, linear regression showed that the MCI progressive group had a significantly higher rate of tOC change over time (mean= 4.54, SD= 11.48) when compared to the MCI stable group (mean = 0.3, SD= 5.54), when adjusting for age, sex and AC score [F(4, 221) = 3.63, P = 0.006] (**Fig 4, panel C**). There was a difference in the rate of change in the first 12 months between the MCI stable and progressive groups (β = 4.144, P = 4.76×10^-07^). survival analysis indicated that for every increase in 3 points of tOC in the first 12 months, the risk of progression from MCI to AD between 12 months and 36 months (in the following two years) increased by 23% (HR = 1.07, 95% CI [1.03,1.08], P = 1.4×10^-04^) (**Fig 4, panel D**).

**Fig. 4.**
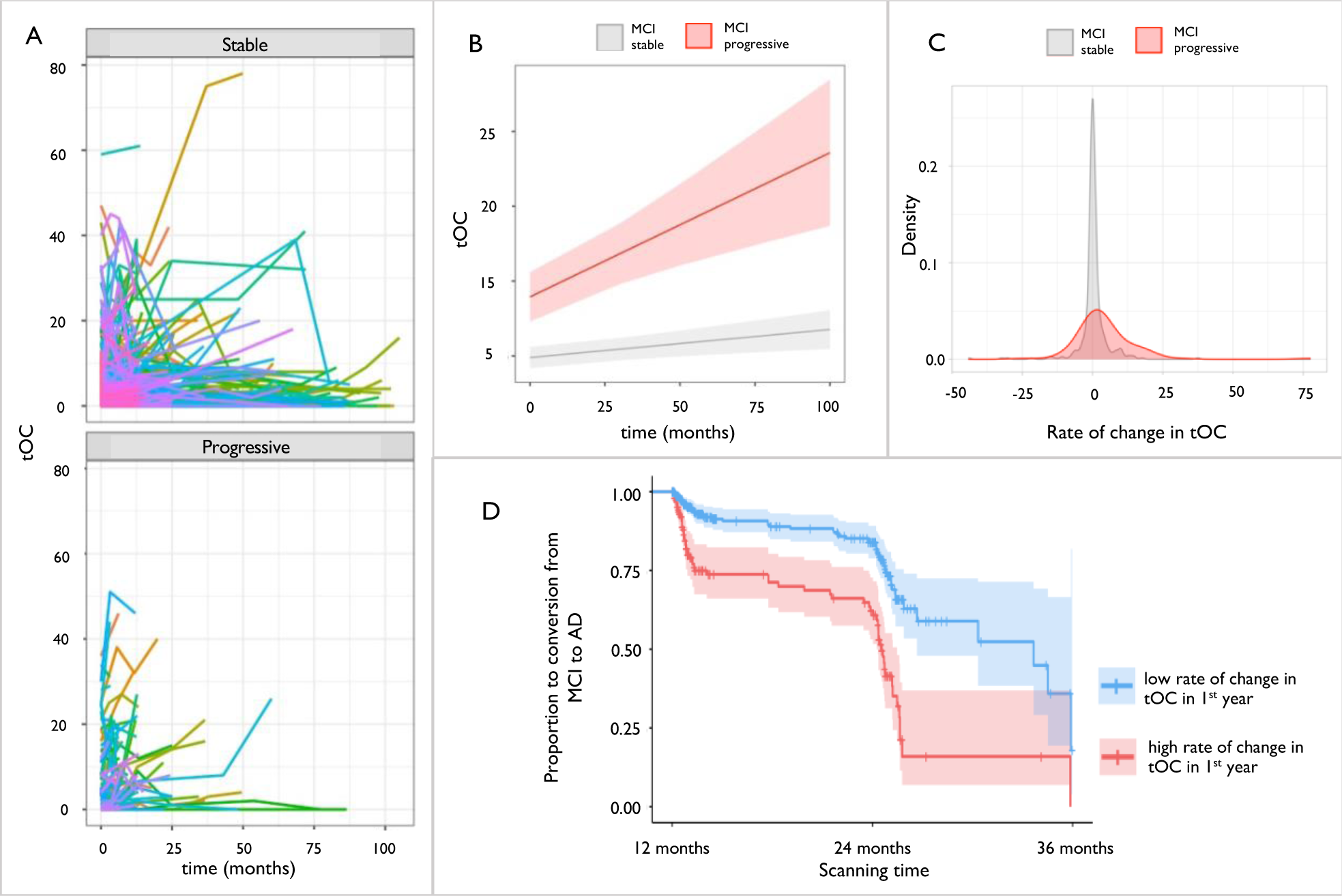
Change in total outlier count (tOC) according to disease conversion status. **(a)** Spaghetti plot of tOC in either MCI stable or MCI progressive. Each line represents an individual change in tOC over the scanning period **(b)** Linear growth model for MCI stable or MCI progressive **(c)** The density spread of the rate of change in tOC in people with MCI. Stable MCI median = 0, MCI progressive = 3 **(d)** Kaplan–Meier plot of MCI progression to AD between 12 and 36 months: the two lines represent a median split of tOC, with <3 classed as low tOC (blue), and ≥3 classed as high tOC (red). Crosses indicate censoring points (i.e., time from baseline at last diagnosis assessment). Filled colour represent the 95% confidence intervals.

### 3.5 Rate of change in tOC correlates with cognitive and amyloid and tau markers

The rate of change of tOC across the whole sample was significantly associated with poorer memory performance [*β* = -1.64, *P* = 1.1×10^-07^] and executive function [*β* = -1.34, *P* = 8.5×10^-07^] in separate linear regression models, controlling for age and sex. To check for the association between the diagnostic group and the two cognitive performance variables, a group-by-memory or group-by-executive function interaction was modelled but was not significant (*P*>0.05) (**Fig.5, panel A;B**). However, the rate of change of tOC across the entire sample was not associated with either a reduction in CSF Aβ 1-42 [*β* = -0.34, *P* = 0.92] or an increase in amyloid PET summary SUV_R_ [*β* = -0.0, *P* = 0.82] when adjusting for age and sex, and neither were influenced by diagnostic group-tOC interaction (*P*>0.05) (**Fig.5, panel C;E**). The rate of change of tOC across the entire sample was significantly associated with an increase in tau PET summary SUV_R_ [*β* = 2.25, *P* = 0.009], but not CSF p-tau181 [*β* = 0.08, *P* = 0.72] when adjusting for age and sex, and neither were influenced by diagnostic group-tOC interaction (*P*>0.05) (**Fig.5, panel D;F**).

**Fig 5:**
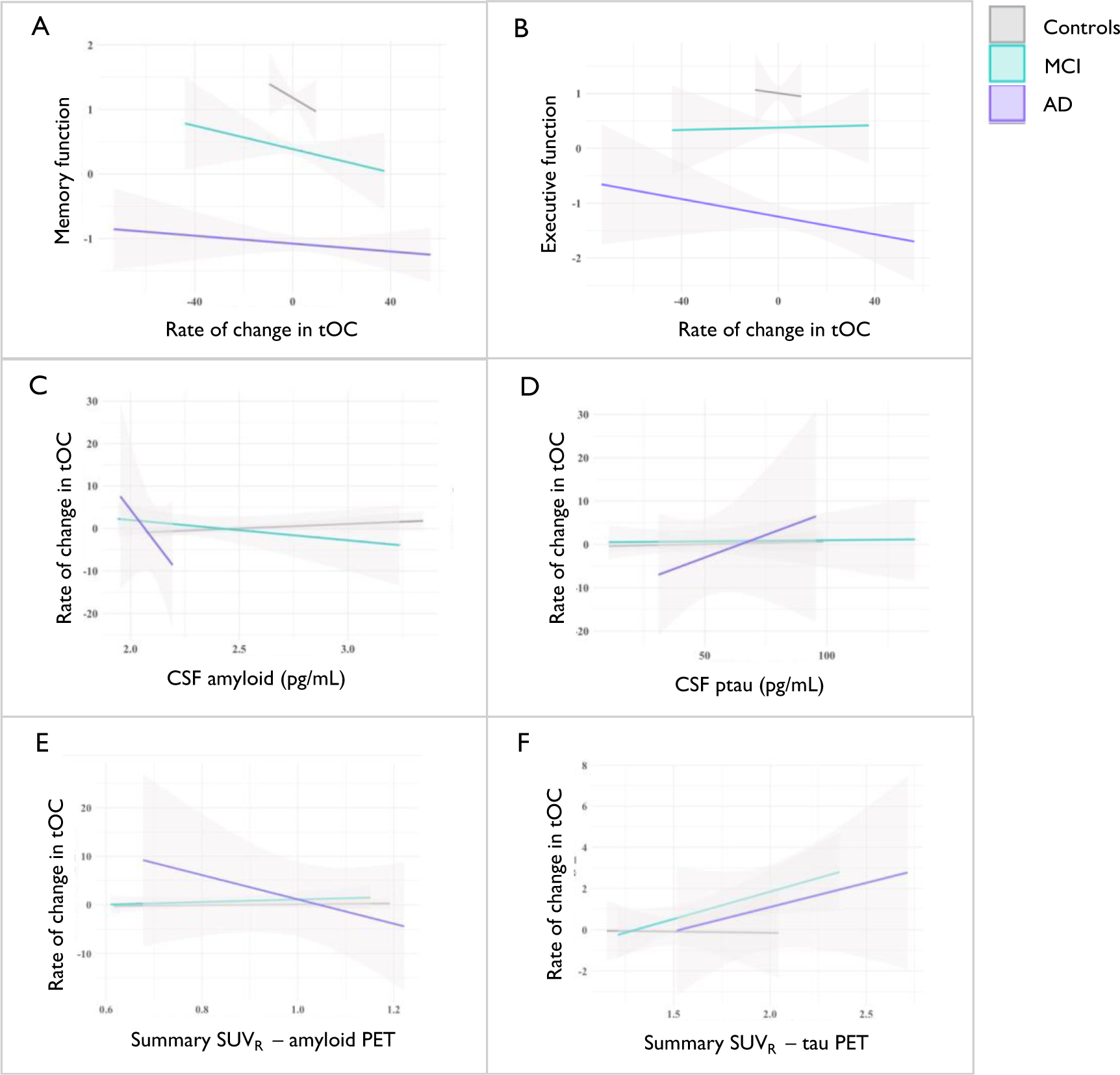
The relationship between cognitive function and CSF markers with the rate of change in tOC. Fitted lines are from a linear regression model per diagnostic group for **(A)** Memory function, **(B)** Executive function, **(C)** CSF Aβ 1-42, and **(D)** CSF phospho-tau **(E)** Summary SUV_R_ amyloid PET **(F)** Summary SUV_R_ tau PET.

### 3.6 The rate of change in tOC is affected by PRS and ApoE ε4 status

An ANOVA adjusting for age and sex showed a difference in the rate of change of tOC according to ApoE ε4 status [*F*(2, 602) = 5.63, *P* = 0.004], which was not influenced by diagnostic group-ApoE ε4 status interaction (*P*>0.05) (**Fig.6, panel A**). This was driven by ApoE ε4 homozygosity (*β* = 2.87, *P*= 0.008), as opposed to ApoE ε4 heterozygosity (*β* = - 0.37, *P*= 0.58).

**Fig. 6.**
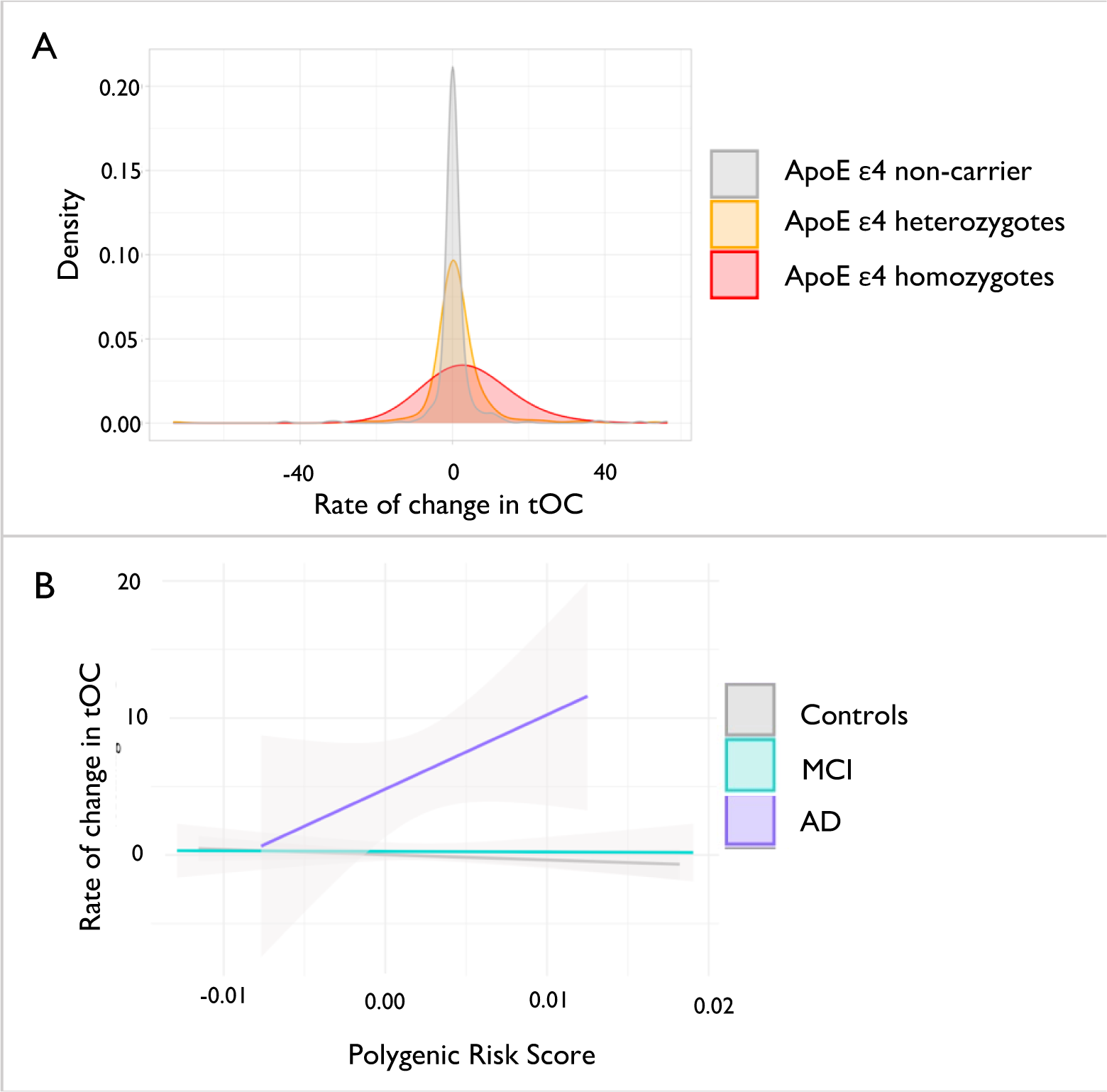
The relationship between genetic risk of AD and the rate of change in tOC. **(a)** The density spread of the rate of change in tOC according to ApoE 4 status **(b)** Linear regression model of Polygenic Risk Score and the rate of change in tOC according to diagnostic group.

Furthermore, when adjusting for age, sex and ApoE ε4 status, linear regression revealed a significant interaction between PRS and the diagnostic group [F(2, 420) = 3.77, P = 0.02], whereby an increase in PRS was driven by the AD group (*β* = 586.2, *P*= 0.01). However, there is no significant relationship between the increase of PRS and the increased rate of change of tOC across all diagnostic groups (*β* = 59.24, *P* = 0.38) (**Fig.6, panel B;C**).

## 4 Discussion

We applied neuroanatomical normative modelling to generate tOC – a personalised brain atrophy marker for patients with AD. This marker is a single summary measure of atrophy across subcortical and cortical brain regions that exceeds expectations. Atrophy variation can also be mapped to an individual brain surface to provide regional information. For the first time, we generate longitudinal tOC and regional outlier measures using serial MRI data acquired up to 9 years (mean follow-up time 2.4 years). We illustrate that neurodegeneration affects different patients with AD in a non-uniform way over the disease course.

A key advantage of neuroanatomical normative modelling is that it provides region-level information by implementing separate normative models for each parcellated neuroanatomical region. In our study, we ran a total of 168 models and from them, we observed that patterns of outliers vary over time in the AD group. For instance, the percentage of participants with AD that had outliers in the hippocampus increased from 47% to 72% in 3 years, suggesting the presence of atrophy in the hippocampus is more heterogenous in earlier stages of the disease and then varies less (i.e., becomes more common) as the disease progresses (**Fig 1**). This is also consistent with our finding that the rate of change in outliers over the study period was highest in the left hippocampus, with 52% of the AD group having marked increasing atrophy in this area (**Fig. 2**). Hippocampal atrophy is seen as characteristic of AD and is included in AD diagnostic criteria, as well as being used in clinical trials and generally considered a key structural marker.^45^ However, our results show that in the ADNI AD sample, 48% of patients do not have greater-than-expected rates of neurodegeneration in that region. Moreover, we found that total MMSE score and or CSF p-tau181 levels were not associated with either having or not having an elevated hippocampal atrophy rate. These results highlight the neuroanatomical heterogeneity in patients with AD, and furthermore reveal what is not always observed in group-average statistical designs. Building on these results, it will be useful to assess the sensitivity of neuroanatomical normative models to the earliest signs of Alzheimer’s in community samples. This would support the future use of the technique to assess MRI in memory clinics and related services, potentially helping to reduce diagnostic uncertainty when assessing patients at risk of Alzheimer’s.

The primary trends of tOC are consistent with previous efforts to quantify neuroanatomical variation in dementia research. Although employing a different neuroanatomical normative modelling technique (hierarchical Bayesian regression ^29^), we previously found that patients with AD had a higher tOC and large inter-individual differences in regional outliers at baseline, when compared to people with MCI and cognitively normal controls.^18, 30^ Here, our longitudinal data showed an increase in tOC in patients with AD. Interestingly, the rate of change in tOC in the AD group increased, suggesting an accelerating accumulation of brain structure outliers over the study period. Therefore, tOC can track neurodegeneration across the disease course in AD (**Fig. 3**).

As well as capturing accumulating atrophy in AD, we show the utility of tOC in the risk disease state of AD, MCI. We show that people with MCI that later progress to AD have increasing rates of tOC (**Fig 4, panels A-C**). This is consistent with previous efforts that have also shown that in a 3-year time period, an increase in 10 tOC points gives a 31.4% chance of clinical progression in 3 years.^30^ Crucially, we further found that the rate of change of tOC over one year (i.e., between baseline scan and 12 month scan), was significantly predictive of progression to dementia in the next 2 years (**Fig 4, panel D**). This could suggest that annual scans for people with MCI to monitor brain health could benefit the clinical decision-making process (e.g., for early AD detection). Moreover, with further validation, it could warrant a run-in period of 12 months in clinical trials, where that have a high or low rate of change in the 1^st^ year can help stratify enrolment.

We also observed that the rate of change in tOC was associated with amyloid and tau PET SUV_R_ and CSF levels (**Fig 5. panels C-F**). This is in line with previous associations of amyloid and tau with neuroanatomical changes in AD,^31–34^ and further supports the established hypothesis that amyloid and tau are risk factors for neurodegeneration,^35^ although it is important to note that the amyloid/tau/neurodegeneration (ATN) interplay is likely to differ from individual to individual.^36, 37^ We also assessed how genetic factors could relate to tOC. We found that an increased rate of tOC in AD is associated with a higher Alzheimer-PRS and ApoE ε4 homozygosity (**Fig 6**), consistent with other structural imaging markers findings.^28, 38–40^ Likewise, our results also support that memory and executive function are risk factors for the accelerated rate of atrophy (**Fig 5. panels A;B**). ^41, 42^ This highlights that individualised measures of neurogenerative change (i.e., tOC) are sensitive to standard CSF, PET, genetic and cognitive AD markers, and may be easily interpreted with current clinical frameworks.^43, 44^

Indeed, accelerated brain atrophy is a widely accepted marker of AD,^4, 46^ yet brain outlier maps and tOC may offer better clinical utility than using raw brain volumes/thicknesses. As neuroanatomical normative modelling generates Z-scores derived from a large reference cohort, they are a standardised value which can allow for individual-level inferences. For instance, in a clinical setting, a patient’s tOC could be compared to another patient’s tOC. Also, the tOC can be derived from serial MRI scans to track atrophy rates across multiple time points. Similar brain atrophy markers derived from normative modelling have already been considered for clinical translation; the Quantitative Neuroradiology Initiative (QNI) has provided a framework to compile reference brain MRI data which can then contextualise a dementia patient’s brain health and provide a personalised score to support the clinical decision-making process eventually.^47^ Building on this idea, our application of neuroanatomical normative modelling can offer regional information on brain health (mapped outlier scores), and improved neuroanatomical normative model estimates by using a large reference cohort.^48^

One setback with translating computational statistical designs in clinical settings are the technical barriers of application and limits to data sharing. However, neuroanatomical normative modelling implementation does not require access to raw scans, as the end-user only requires a pre-trained reference model, which contains no identifiable data. Furthermore, scripts to generate individual Z-scores, tOC and outlier maps are openly available.^49^

Neuroanatomical normative modelling also has the potential to aid in trials of AD therapeutics. For example, it could be used to stratify people for trial enrolment based on the extent (tOC) and spatial distribution of their brain atrophy, to identify subgroups based on different atrophy patterns or as a personalised outcome measure, where the impact of the treatment using unique brain ‘fingerprints’ can be quantified, increasing power and sensitivity to subtle changes over time.^50^

Yet before clinical and drug trial implementation, additional diversification of datasets is needed. Although our reference (training) dataset is large, it is over-representative of European ancestry due to the datasets predominantly from academic studies (which do not match either regional or global population demographics). ^51, 52^ Though ADNI participants are mostly European-ancestry ^53^, caution should be made when transferring the model to diverse datasets, or participants from underrepresented demographics.^54^ Future work will require the reference dataset and clinical datasets to include participants from non-research studies, different social-economic backgrounds and ethnicities to reduce bias and mitigate healthcare inequalities.^55^

Moreover, further optimisations of neuroanatomical normative modelling are possible. Although scanner effects and non-gaussian nature of MRI data were accounted for, unwanted noise might be generated by longitudinal data collection. To assess this, it will be useful to understand test-retest reliability by calculating the difference in scans which have been acquired in close succession (< one week). Also, unwanted noise might be limited at the stage of data processing by implementing the longitudinal FreeSurfer pipeline.^56^ Moreover, in our model, brain regions are treated independently, yet is likely that regional Z-score are inter-correlated, particularly between neighbouring or bilateral regions. Solutions to this would be to consider the spatial extent of affected voxels and the magnitude in those voxels, ^57^ and to employ normative models which use brain connectivity data, which have shown recent promise. ^51^

To conclude, we show that atrophy across MCI and AD disease course differs at the individual level – and that this can be tracked using tOC and brain Z-score maps generated by neuroanatomical normative modelling. Our studies further corroborate the utility of tOC and brain Z-score maps as personalised brain atrophy markers for patients with AD and predict disease progression in people with MCI. The next steps are to diversify the training and clinical data used in neuroanatomical normative modelling for AD research, in efforts to validate these markers for future translation into clinical settings and in drug trial design.

## Data Availability

ADNI data used in this study are publicly available and can be requested following ADNI Data Sharing and Publications Committee guidelines: http://adni.loni.usc.edu/data-samples/access-data/ Reference data are available online at: https://github.com/predictive-clinical-neuroscience/braincharts The neuroanatomical normative model was generated using the PCNtoolkit software package: https://github.com/amarquand/PCNtoolkit).

## Acknowledgements

Data collection and sharing for this project was funded by ADNI (National Institutes of Health Grant U01 AG024904) and DOD ADNI (Department of Defence award number W81XWH-12-2-0012). ADNI is funded by the National Institute on Aging, the National Institute of Biomedical Imaging and Bioengineering, and through generous contributions from the following: AbbVie, Alzheimer’s Association; Alzheimer’s Drug Discovery Foundation; Araclon Biotech; BioClinica, Inc.; Biogen; Bristol-Myers Squibb Company; CereSpir, Inc.; Cogstate; Eisai Inc.; Elan Pharmaceuticals, Inc.; Eli Lilly and Company; EuroImmun; F. Hoffmann-La Roche Ltd and its affiliated company Genentech, Inc.; Fujirebio; GE Healthcare; IXICO Ltd.;Janssen Alzheimer Immunotherapy Research & Development, LLC.; Johnson & Johnson Pharmaceutical Research & Development LLC.; Lumosity; Lundbeck; Merck & Co., Inc.;Meso Scale Diagnostics, LLC.; NeuroRx Research; Neurotrack Technologies; Novartis Pharmaceuticals Corporation; Pfizer Inc.; Piramal Imaging; Servier; Takeda Pharmaceutical Company; and Transition Therapeutics. The Canadian Institutes of Health Research is providing funds to support ADNI clinical sites in Canada. Private sector contributions are facilitated by the Foundation for the National Institutes of Health (www.fnih.org). The grantee organization is the Northern California Institute for Research and Education, and the study is coordinated by the Alzheimer’s Therapeutic Research Institute at the University of Southern California. ADNI data are disseminated by the Laboratory for Neuro Imaging at the University of Southern California.

## Present/permanent address

Centre for Medical Image Computing, Medical Physics and Biomedical Engineering, University College London, London, UK

## Research in Context

### Systematic review

The authors reviewed the literature using traditional (e.g., PubMed) sources. AD atrophy is heterogenous at the individual level throughout the disease course – neuroanatomical normative modelling is a potential quantitative technique to capture this. Relevant literature is cited.

### Interpretation

Neuroanatomical normative modelling can generate personalised brain atrophy markers which can map changes over time in AD. We illustrate that neurodegeneration rates are heterogenous over the disease course and reveal what cannot be seen using traditional group average statistics. This is consistent with previous AD and normative modelling research.

### Future directions

Further validation of our personalised brain atrophy marker could be conducted in community-based samples, that comprise patients with both earlier and late-stage AD (to capture the full disease course). Future studies should assess if these individualised markers are sensitive to (1) capturing the deceleration of atrophy in disease-modifying clinical trials (2) being implemented as a decision-making tool in clinical settings.

## References

1. Aisen PS, Cummings J, Jack CR, et al. On the path to 2025: Understanding the Alzheimer’s disease continuum. Alzheimers Res Ther. 2017;9(1):1–10. doi:10.1186/S13195-017-0283-5/TABLES/2

2. Eid A, Mhatre I, Richardson JR. Gene-environment interactions in Alzheimer’s Disease: A potential path to precision medicine. Pharmacol Ther. 2019;199:173. doi:10.1016/J.PHARMTHERA.2019.03.005

3. Verdi S, Marquand AF, Schott JM, Cole JH. Beyond the average patient: how neuroimaging models can address heterogeneity in dementia. Brain. 2021;144(10):2946–2953. doi:10.1093/BRAIN/AWAB165

4. Whitwell JL. Progression of atrophy in Alzheimer’s disease and related disorders. Neurotox Res. 2010;18(3-4):339–346. doi:10.1007/S12640-010-9175-1

5. Gauthier S, Albert M, Fox N, et al. Why has therapy development for dementia failed in the last two decades? Alzheimer’s and Dementia. 2016;12(1):60–64. doi:10.1016/j.jalz.2015.12.003

6. Cummings J, Feldman HH, Scheltens P. The “rights” of precision drug development for Alzheimer’s disease. Alzheimers Res Ther. 2019;11(1). doi:10.1186/s13195-019-0529-5

7. Aisen PS, Cummings J, Jack CR, et al. On the path to 2025: Understanding the Alzheimer’s disease continuum. Alzheimers Res Ther. 2017;9(1). doi:10.1186/s13195-017-0283-5

8. Ryan J, Fransquet P, Wrigglesworth J, Lacaze P. Phenotypic heterogeneity in dementia: A challenge for epidemiology and biomarker studies. Front Public Health. 2018;6. doi:10.3389/fpubh.2018.00181

9. Duara R, Barker W. Heterogeneity in Alzheimer’s Disease Diagnosis and Progression Rates: Implications for Therapeutic Trials. Neurotherapeutics 2022 19:1. 2022;19(1):8–25. doi:10.1007/S13311-022-01185-Z

10. Devi G, Scheltens P. Heterogeneity of Alzheimer’s disease: Consequence for drug trials? Alzheimers Res Ther. 2018;10(1):1–3. doi:10.1186/S13195-018-0455-Y/METRICS

11. Marquand AF, Wolfers T, Mennes M, Buitelaar J, Beckmann CF. Beyond Lumping and Splitting: A Review of Computational Approaches for Stratifying Psychiatric Disorders. Biol Psychiatry Cogn Neurosci Neuroimaging. 2016;1(5):433–447. doi:10.1016/j.bpsc.2016.04.002

12. ten Kate M, Dicks E, Visser PJ, et al. Atrophy subtypes in prodromal Alzheimer’s disease are associated with cognitive decline. Brain. 2018;141(12):3443–3456. doi:10.1093/brain/awy264

13. Dementia. Dementia: Assessment, management and support for people living with dementia and their carers. Published online 2018. Accessed February 10, 2022. https://www.ncbi.nlm.nih.gov/books/NBK513207/

14. Rutherford S, Fraza C, Dinga R, et al. Charting brain growth and aging at high spatial precision. Elife. 2022;11. doi:10.7554/ELIFE.72904

15. Marquand AF, Kia SM, Zabihi M, Wolfers T, Buitelaar JK, Beckmann CF. Conceptualizing mental disorders as deviations from normative functioning. Mol Psychiatry. 2019;24(10):1415–1424. doi:10.1038/s41380-019-0441-1

16. Cole TJ. The development of growth references and growth charts. Ann Hum Biol. 2012;39(5):382–394. doi:10.3109/03014460.2012.694475

17. Verdi S, Kia SM, Yong KXX, et al. Revealing Individual Neuroanatomical Heterogeneity in Alzheimer Disease Using Neuroanatomical Normative Modeling. Neurology. Published online May 1, 2023:10.1212/WNL.0000000000207298. doi:10.1212/WNL.0000000000207298

18. Loreto F, Verdi S, Kia SM, et al. Examining real-world Alzheimer’s disease heterogeneity using neuroanatomical normative modelling. medRxiv. Published online November 4, 2022:2022.11.02.22281597. doi:10.1101/2022.11.02.22281597

19. Kia S, Huijsdens H, Rutherford S, et al. Federated Multi-Site Normative Modeling using Hierarchical Bayesian Regression. Published online 2021. doi:10.1101/2021.05.28.446120

20. Jack CR, Bernstein MA, Borowski BJ, et al. Update on the Magnetic Resonance Imaging core of the Alzheimer’s Disease Neuroimaging Initiative. Alzheimer’s & Dementia. 2010;6(3):212–220. doi:10.1016/J.JALZ.2010.03.004

21. FreeSurferVersion3 - Free Surfer Wiki. Accessed May 27, 2023. https://surfer.nmr.mgh.harvard.edu/fswiki/FreeSurferVersion3#AsegAtlas

22. Fischl B. FreeSurfer. Neuroimage. Published online 2012. doi:10.1016/j.neuroimage.2012.01.021

23. Fraza CJ, Dinga R, Beckmann CF, Marquand AF. Warped Bayesian linear regression for normative modelling of big data. Neuroimage. 2021;245:118715. doi:10.1016/J.NEUROIMAGE.2021.118715

24. Raket LL, Palmqvist S, Mattsson-Carlgren N, Hansson O, Nordisk N, Søborg D. An amyloid-cognition composite score for estimating time-consistent biomarker trajectories across the Alzheimer’s continuum. Alzheimer’s & Dementia. 2022;18(S6):e061629. doi:10.1002/ALZ.061629

25. Kühnel L, Berger AK, Markussen B, Raket LL. Simultaneous modeling of Alzheimer’s disease progression via multiple cognitive scales. Stat Med. 2021;40(14):3251–3266. doi:10.1002/SIM.8932

26. Raket LL. Statistical Disease Progression Modeling in Alzheimer Disease. Front Big Data. 2020;3:553735. doi:10.3389/FDATA.2020.00024/BIBTEX

27. Landau SM, Breault C, Joshi AD, et al. Amyloid-β imaging with Pittsburgh compound B and florbetapir: comparing radiotracers and quantification methods. J Nucl Med. 2013;54(1):70–77. doi:10.2967/JNUMED.112.109009

28. Altmann A, Scelsi MA, Shoai M, et al. A comprehensive analysis of methods for assessing polygenic burden on Alzheimer’s disease pathology and risk beyond APOE. Brain Commun. 2020;2(1). doi:10.1093/BRAINCOMMS/FCZ047

29. Kia SM, Huijsdens H, Rutherford S, et al. Closing the life-cycle of normative modeling using federated hierarchical Bayesian regression. PLoS One. 2022;17(12). doi:10.1371/JOURNAL.PONE.0278776

30. Verdi S, Kia SM, Yong K, et al. Revealing Individual Neuroanatomical Heterogeneity in Alzheimer’s Disease. medRxiv. Published online July 3, 2022:2022.06.30.22277053. doi:10.1101/2022.06.30.22277053

31. Tosun D, Schuff N, Shaw LM, Trojanowski JQ, Weiner MW. Relationship Between CSF Biomarkers of Alzheimer’s Disease and Rates of Regional Cortical Thinning in ADNI Data. J Alzheimers Dis. 2011;26(0 3):77. doi:10.3233/JAD-2011-0006

32. Gordon BA, McCullough A, Mishra S, et al. Cross-sectional and longitudinal atrophy is preferentially associated with tau rather than amyloid β positron emission tomography pathology. Alzheimer’s & Dementia: Diagnosis, Assessment & Disease Monitoring. 2018;10:245–252. doi:10.1016/J.DADM.2018.02.003

33. Mohanty R, Ferreira D, Nordberg A, Westman E. Associations between different tau-PET patterns and longitudinal atrophy in the Alzheimer’s disease continuum: biological and methodological perspectives from disease heterogeneity. Alzheimers Res Ther. 2023;15(1):1–16. doi:10.1186/S13195-023-01173-1/FIGURES/5

34. Joie R La, Visani A V., Baker SL, et al. Prospective longitudinal atrophy in Alzheimer’s disease correlates with the intensity and topography of baseline tau-PET. Sci Transl Med. 2020;12(524). doi:10.1126/SCITRANSLMED.AAU5732

35. Bloom GS. Amyloid-β and tau: the trigger and bullet in Alzheimer disease pathogenesis. JAMA Neurol. 2014;71(4):505–508. doi:10.1001/JAMANEUROL.2013.5847

36. Grøntvedt GR, Lauridsen C, Berge G, et al. The Amyloid, Tau, and Neurodegeneration (A/T/N) Classification Applied to a Clinical Research Cohort with Long-Term Follow-Up. Journal of Alzheimer’s Disease. 2020;74(3):829. doi:10.3233/JAD-191227

37. Hadjichrysanthou C, Evans S, Bajaj S, Siakallis LC, McRae-Mckee K, De Wolf F. The dynamics of biomarkers across the clinical spectrum of Alzheimer’s disease. Alzheimers Res Ther. 2020;12(1):1–16. doi:10.1186/S13195-020-00636-Z/FIGURES/5

38. Harrison JR, Mistry S, Muskett N, Escott-Price V, Brookes K. From Polygenic Scores to Precision Medicine in Alzheimer’s Disease: A Systematic Review. Journal of Alzheimer’s Disease. 2020;74(4):1271. doi:10.3233/JAD-191233

39. Emrani S, Arain HA, DeMarshall C, Nuriel T. APOE4 is associated with cognitive and pathological heterogeneity in patients with Alzheimer’s disease: a systematic review. Alzheimer’s Research & Therapy 2020 12:1. 2020;12(1):1–19. doi:10.1186/S13195-020-00712-4

40. Mishra S, Blazey TM, Holtzman DM, et al. Longitudinal brain imaging in preclinical Alzheimer disease: impact of APOE ε4 genotype. Brain. 2018;141(6):1828–1839. doi:10.1093/BRAIN/AWY103

41. Jutten RJ, Sikkes SAM, Van der Flier WM, Scheltens P, Visser PJ, Tijms BM. Finding Treatment Effects in Alzheimer Trials in the Face of Disease Progression Heterogeneity. Neurology. 2021;96(22):e2673–e2684. doi:10.1212/WNL.0000000000012022

42. Sintini I, Graff-Radford J, Senjem ML, et al. Longitudinal neuroimaging biomarkers differ across Alzheimer’s disease phenotypes. Brain. 2020;143(7):2281–2294. doi:10.1093/BRAIN/AWAA155

43. Jack CR, Bennett DA, Blennow K, et al. NIA-AA Research Framework: Toward a biological definition of Alzheimer’s disease. Alzheimer’s & Dementia. 2018;14(4):535–562. doi:10.1016/J.JALZ.2018.02.018

44. Boccardi M, Dodich A, Albanese E, et al. The strategic biomarker roadmap for the validation of Alzheimer’s diagnostic biomarkers: methodological update. Eur J Nucl Med Mol Imaging. 2021;48(7):2070–2085. doi:10.1007/S00259-020-05120-2/FIGURES/1

45. McKhann GM, Knopman DS, Chertkow H, et al. The diagnosis of dementia due to Alzheimer’s disease: Recommendations from the National Institute on Aging-Alzheimer’s Association workgroups on diagnostic guidelines for Alzheimer’s disease. Alzheimer’s & Dementia. 2011;7(3):263–269. doi:10.1016/J.JALZ.2011.03.005

46. Leung KK, Bartlett JW, Barnes J, Manning EN, Ourselin S, Fox NC. Cerebral atrophy in mild cognitive impairment and Alzheimer disease. Neurology. 2013;80(7):648–654. doi:10.1212/WNL.0B013E318281CCD3

47. Goodkin O, Pemberton H, Vos SB, et al. The quantitative neuroradiology initiative framework: Application to dementia. British Journal of Radiology. Published online 2019. doi:10.1259/bjr.20190365

48. Bozek J, Griffanti L, Lau S, Jenkinson M. Normative models for neuroimaging markers: Impact of model selection, sample size and evaluation criteria. Neuroimage. 2023;268:119864. doi:10.1016/J.NEUROIMAGE.2023.119864

49. GitHub - predictive-clinical-neuroscience/braincharts. Accessed April 30, 2023. https://github.com/predictive-clinical-neuroscience/braincharts

50. Duara R, Barker W. Heterogeneity in Alzheimer’s Disease Diagnosis and Progression Rates: Implications for Therapeutic Trials. Neurotherapeutics 2022 19:1. 2022;19(1):8–25. doi:10.1007/S13311-022-01185-Z

51. Rutherford S, Barkema P, Tso IF, et al. Evidence for embracing normative modeling. Elife. 2023;12. doi:10.7554/ELIFE.85082

52. Benkarim O, Paquola C, Park BY, et al. Population heterogeneity in clinical cohorts affects the predictive accuracy of brain imaging. PLoS Biol. 2022;20(4). doi:10.1371/JOURNAL.PBIO.3001627

53. Ashford MT, Raman R, Miller G, et al. Screening and enrollment of underrepresented ethnocultural and educational populations in the Alzheimer’s Disease Neuroimaging Initiative (ADNI). Alzheimer’s & Dementia. 2022;18(12):2603–2613. doi:10.1002/ALZ.12640

54. Weiner MW, Veitch DP, Miller MJ, et al. Increasing participant diversity in AD research: Plans for digital screening, blood testing, and a community-engaged approach in the Alzheimer’s Disease Neuroimaging Initiative 4. Alzheimer’s & Dementia. 2023;19(1):307–317. doi:10.1002/ALZ.12797

55. Hampel H, Au R, Mattke S, et al. Designing the next-generation clinical care pathway for Alzheimer’s disease. Nature Aging 2022 2:8. 2022;2(8):692-703. doi:10.1038/s43587-022-00269-x

56. Hedges EP, Dimitrov M, Zahid U, et al. Reliability of structural MRI measurements: The effects of scan session, head tilt, inter-scan interval, acquisition sequence, FreeSurfer version and processing stream. Neuroimage. 2022;246:118751. doi:10.1016/J.NEUROIMAGE.2021.118751

57. Smith SM, Nichols TE. Threshold-free cluster enhancement: Addressing problems of smoothing, threshold dependence and localisation in cluster inference. Neuroimage. Published online 2009. doi:10.1016/j.neuroimage.2008.03.061

